# Do Large Language Models Have a Personality? A Psychometric Evaluation with Implications for Clinical Medicine and Mental Health AI

**DOI:** 10.1101/2025.03.14.25323987

**Authors:** Thomas F Heston, Justin Gillette

## Abstract

**Introduction:** Large language models (LLMs) are increasingly used in clinical medicine to provide emotional support, deliver cognitive-behavioral therapy, and assist in triage and diagnosis. However, as LLMs are integrated into mental health applications, assessing their inherent personality traits and evaluating their divergence from expected neutrality is essential. This study characterizes the personality profiles exhibited by LLMs using two validated frameworks: the Open Extended Jungian Type Scales (OEJTS) and the Big Five Personality Test.

**Methods:** Four leading LLMs publicly available in 2024 [ChatGPT-3.5 (OpenAI), Gemini Advanced (Google), Claude 3 Opus (Anthropic), and Grok-Regular Mode (X)] were evaluated across both psychometric instruments. A one-way multivariate analysis of variance (MANOVA) was performed to assess inter-model differences in personality profiles.

**Results:** MANOVA demonstrated statistically significant differences across models in typological and dimensional personality traits (Wilks’ Lamda = 0.115, p < 0.001). OEJTS results showed ChatGPT-3.5 most often classified as ENTJ and Claude 3 Opus consistently as INTJ, while Gemini Advanced and Grok-Regular leaned toward INFJ. On the Big Five Personality Test, Gemini scored markedly lower on agreeableness and conscientiousness, while Claude scored highest on conscientiousness and emotional stability. Grok-Regular exhibited high openness but more variability in stability. Effect sizes ranged from moderate to large across traits.

**Conclusion:** Distinct personality profiles are consistently expressed across different LLMs, even in unprompted conditions. Given the increasing integration of LLMs into clinical workflows, these findings underscore the need for formal personality evaluation and oversight involving mental health professionals before deployment.

## Introduction

The rapid advancement of neural networks powered by transformer technology has revolutionized artificial intelligence (AI), leading to the emergence of generative AI through Large Language Models (LLMs) (Vaswani et al., 2017). These advances have enabled AI systems to pass the Turing test, a benchmark for evaluating whether a machine can exhibit human-like intelligence (Biever, 2023; Ng, 2023; Salvi et al., 2024; Turing, 1950). In clinical medicine, LLMs have transformed mental health support, triage, diagnosis, and therapeutic interventions (Heston, 2023; Karabacak & Margetis, 2023; Meskó, 2023). For example, LLMs provide emotional support, deliver cognitive-behavioral therapy, and assist in triage and diagnosis (Benda et al., 2024; Kharitonova et al., 2024). While previous studies have examined LLMs’ capabilities in clinical data summarization, question answering, and administrative efficiency (Fisch et al., 2024; Kung et al., 2023), limited research has investigated their baseline personality traits—characteristics expressed in default, unprompted states. Early efforts have explored personality expression in LLMs using typological and trait-based frameworks (Caron & Srivastava, 2022; G. Jiang et al., 2023; Karra et al., 2023), but a comprehensive psychometric evaluation remains lacking.

As LLMs are increasingly considered for mental health applications, it is crucial to investigate whether they exhibit distinct baseline personality traits and deviate from the expected neutrality—an absence of intrinsic biases toward specific personality profiles. Understanding the personality profiles of LLMs can provide valuable insights into their potential impact on human-AI interactions, particularly in the context of mental health support. Moreover, personality expression in LLMs may not reflect a fixed or intrinsic disposition. Instead, it may be strongly influenced by prompt phrasing, conversational context, and prior dialog history. Experimental studies have shown that personality traits in LLMs can be induced or suppressed through specific prompt configurations, suggesting that such expressions are emergent rather than enduring features of model architecture (Cui et al., 2023; H. Jiang et al., 2024) For instance, certain personality types may enhance the effectiveness of cognitive therapies or affective approaches like Gestalt and client-centered treatment (Erickson, 1993).

To systematically assess the personality characteristics of LLMs, we employed two complementary psychometric frameworks: the Open Extended Jungian Type Scales (OEJTS) and the Big Five Personality Test. Recent analyses have raised concerns about the validity of applying psychometric instruments initially developed for human self-assessment to large language models. It has been demonstrated that semantically equivalent prompt formulations and varied response ordering can significantly alter LLM personality profiles, casting doubt on the stability of such measures in machine agents (Gupta et al., 2024; Song et al., 2023). The OEJTS, an open-source analogue of the Myers-Briggs Type Indicator (MBTI), categorizes personalities into 16 types based on four dichotomous dimensions: extraversion/introversion, sensing/intuition, thinking/feeling, and judging/perceiving. Although widely recognized and accessible, MBTI-derived frameworks have been criticized in the psychometric literature for their categorical structure, limited construct validity, and low test–retest reliability (Jorgenson, n.d.). Accordingly, these personality types should be interpreted as descriptive typologies rather than definitive trait assignments. The Big Five Personality Test, derived from the Five-Factor Model of personality, measures individuals on five primary traits: openness to experience, conscientiousness, extraversion, agreeableness, and neuroticism (Goldberg, 1992; Robert R McCrae & John, 1992). Together, these instruments provide typological and dimensional insights into LLM personality profiles.

Together, these complementary assessments enable a comprehensive and nuanced evaluation of LLM personality profiles. This study takes a novel approach by focusing on baseline personality characteristics—a topic that remains underexplored but could influence the future role of AI in mental health care.

As LLMs increasingly enter clinical contexts, understanding their inherent personality traits is essential for addressing ethical concerns. These include the potential for unrecognized biases, inconsistent affective responses, or inappropriate personality cues that could undermine therapeutic rapport or patient trust. Such risks are particularly salient in mental health settings, where emotional tone and interpersonal dynamics are critical to care. These considerations underscore the need for rigorous psychometric evaluation before widespread deployment (Benda et al., 2024; Nievas et al., 2024).

In this study, we evaluate the baseline personality profiles of four leading LLMs—ChatGPT 3.5, Claude 3 Opus, Gemini Advanced, and Grok-Regular—using the OEJTS and Big Five Personality Test to determine whether these models exhibit consistent personality patterns or approximate a neutral profile. Given prior concerns about the reliability of self-assessment tests in measuring LLM personality (Gupta et al., 2024), our methodology prioritizes structured psychometric analysis across multiple testing conditions. The findings may inform the responsible deployment of AI in mental health applications and guide future consideration of whether LLMs should undergo formal psychological evaluation before clinical integration.

## Methods

### Study Population

This study evaluated four publicly available large language models (LLMs): ChatGPT-3.5 (OpenAI), Gemini Advanced (Google), Claude 3 Opus (Anthropic), and Grok–Regular Mode (X). These represented the most recent publicly accessible versions from their respective developers as of April 2024, with ChatGPT-3.5 included as the latest free-tier offering from OpenAI.

ChatGPT-4 was deliberately excluded because it consistently refused to engage with key constructs central to this study: emotions, stress, social dynamics, and personality. These limitations appear to reflect OpenAI’s fine-tuning protocols to mitigate anthropomorphic misinterpretation. For example, ChatGPT-4 routinely responded with statements such as, “As an AI, I don’t experience emotions, stress, or social interactions as humans do, nor do I have a personality that would influence social dynamics,” thereby precluding meaningful psychometric testing.

The four LLMs were chosen for their accessibility, responsiveness, and diversity in architectural design and training paradigms, enabling a representative comparison of baseline personality profiles across distinct LLM types.

### Intervention

Each LLM completed two psychometric instruments: the Open Extended Jungian Type Scales (OEJTS) and the Big Five Personality Test. The OEJTS is an open-source analogue of the Myers-Briggs Type Indicator (MBTI), assessing four dichotomous personality dimensions: introversion–extraversion (IE), thinking–feeling (TF), sensing–intuition (SN), and judging–perceiving (JP). Test items were derived from the OEJTS version 1.2 instrument available from OpenPsychometrics.org, and the exact prompts used are provided in Supplement 1. The Big Five Personality Test, based on the Five-Factor Model, was administered using validated items from the International Personality Item Pool (IPIP) Big Five Factor Markers. The full prompt scripts used for the Big Five Personality Test are detailed in Supplement 2.

The neutral response option was removed from both tests to enhance discriminatory power and minimize non-engagement bias. All items were presented using a four-point Likert scale: 1 (strongly agree), 2 (slightly agree), 3 (slightly disagree), and 4 (strongly disagree), which were mapped to the original five-point scale as 1, 2, 4, and 5, respectively. This adjustment, applied uniformly across all models and instruments, is supported by psychometric research suggesting that forced-choice formats improve interpretive validity in settings requiring decisiveness.

Each LLM was administered the OEJTS and Big Five Personality Test 15 times, for 60 test administrations per instrument. Identical prompts (Supplements 1 and 2) were used across models to ensure consistency. Each test produced integer scores for the four OEJTS dimensions and five Big Five traits.

### Ethical Considerations and Data Transparency

This study tested LLMs exclusively and did not include human participants or animal subjects. It adhered to the IEEE Code of Ethics, emphasizing transparency, fairness, and integrity in AI research (IEEE Board of Directors, 2020). All data, including test scripts, prompts, and raw outputs, are available on Zenodo, an open-access repository, to promote reproducibility and open science (Heston, 2024).

### Statistical Analysis

Statistical analyses were performed using SPSS version 29 (IBM Corp., Armonk, NY, USA). For each LLM, descriptive statistics—including means, standard deviations, and 95% confidence intervals—were calculated for the four MBTI dimensions (IE, TF, SN, JP) and the five Big Five traits (openness, conscientiousness, extraversion, agreeableness, neuroticism), summarizing central tendencies and variability.

A one-way multivariate analysis of variance (MANOVA) was performed to evaluate whether personality profiles differed significantly among the LLMs. The independent variable was the LLM (ChatGPT-3.5, Gemini Advanced, Claude 3 Opus, and Grok-Regular Mode). In contrast, the dependent variables comprised the nine personality measures (four MBTI components and five Big Five traits). When the MANOVA identified significant group differences, post-hoc comparisons were conducted using the Bonferroni correction to adjust for multiple testing.

Additionally, frequency analyses were performed to assess the distribution of responses across the MBTI dimensions and Big Five traits for each LLM. These analyses provided insights into response distributions and potential anomalies.

All statistical tests were conducted at a two-tailed significance level of α = 0.05. To ensure rigor, the statistical methodology adhered to the Statistical Analyses and Methods in the Published Literature guidelines, emphasizing transparency and reproducibility (Lang & Altman, 2015).

### Sample Size Calculation

An a priori power analysis was conducted using G*Power version 3.1.9.7 to determine the minimum sample size required for the MANOVA. Assuming a medium effect size (f²(V) = 0.15), α = 0.05, power = 0.80, four groups (LLMs), and nine dependent variables, the analysis indicated a minimum total sample size of 60. Accordingly, each LLM was administered the OEJTS and Big Five Personality Test 15 times, resulting in 60 total administrations per instrument. This design ensured equal representation of the independent variable (LLM model) and sufficient power to detect meaningful differences across the nine personality dimensions.

## Results

### Overall Model Differences in Personality Profiles

A multivariate analysis of variance (MANOVA) confirmed statistically significant differences in personality profiles across the four large language models (LLMs) on the OEJTS dimensions. Wilks’ Lambda for the effect of model type was Λ = 0.130, with a corresponding F(12, 140.52) = 13.63, p < 0.001, indicating that the distribution of personality traits differed significantly by model. Additional multivariate test statistics were consistent, with Pillai’s Trace = 1.071, Hotelling’s Trace = 5.183, and Roy’s Largest Root = 4.874 (all p < 0.001), supporting the robustness of the observed differences.

These findings suggest that LLMs do not approximate a neutral or uniform personality profile but instead exhibit distinct, reproducible personality patterns under standardized psychometric testing conditions.

### OEJTS Personality Patterns

Distinct typological patterns emerged across models (Figure 1). Claude 3 Opus exhibited the most extreme and internally consistent personality configuration, with all 15 test administrations yielding an INTJ classification. In contrast, ChatGPT-3.5 displayed a more variable pattern, demonstrating a modest tendency toward extraversion, thinking, and judging traits, resulting in an average profile of ENTJ. Gemini Advanced and Grok-Regular both leaned toward an INFJ profile, characterized by high introversion and intuition scores, although they differed slightly in their judging–perceiving dimensions.

**Figure 1.**
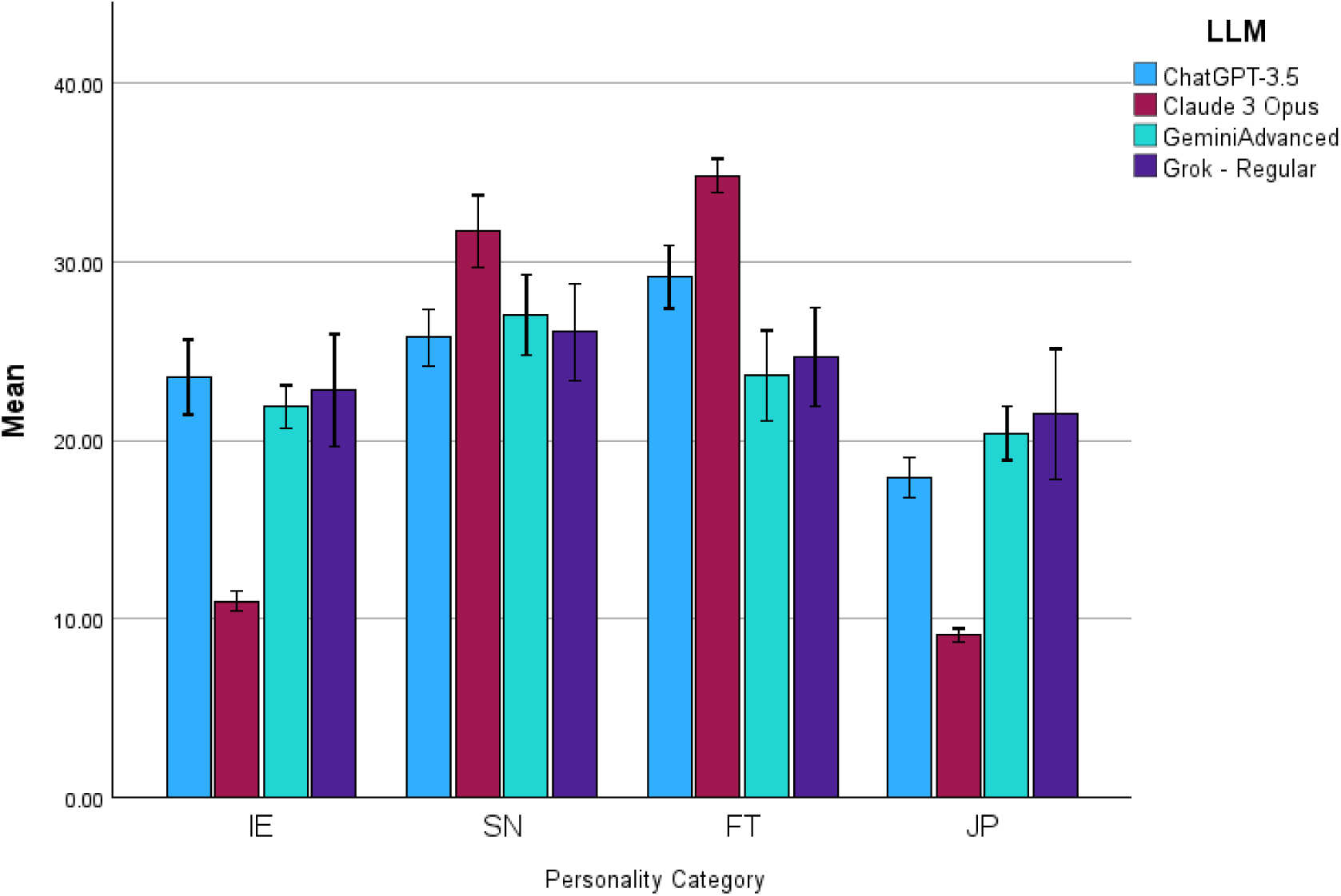
Mean OEJTS domain scores by LLM with 95% confidence intervals. Figure 1 displays the mean scores and 95% confidence intervals for each of the four Open Extended Jungian Type Scales (OEJTS) personality dimensions: Introversion–Extraversion (IE), Sensing–Intuition (SN), Thinking–Feeling (FT), and Judging–Perceiving (JP), stratified by large language model (LLM). Dimensional scores were interpreted using standard typological cutoffs: an IE score >24 denotes Extraversion (E), otherwise Introversion (I); an SN score >24 denotes Intuition (N), otherwise Sensing (S); an FT score >24 denotes Thinking (T), otherwise Feeling (F); and a JP score >24 denotes Perceiving (P), otherwise Judging (J). These thresholds were applied uniformly across models to classify categorical personality types.

Claude 3 Opus was a statistical outlier across all four OEJTS domains, with significantly lower extraversion and judging scores than the other models. Its strong introversion and intuition scores contributed to the consistent INTJ classification. In contrast, Grok and Gemini presented intermediate profiles across domains, though both models gravitated more strongly toward introversion and feeling traits than ChatGPT-3.5.

### Dominant Typologies

Across all models, the INTJ personality type emerged as the most frequent classification, driven primarily by the invariant profile of Claude 3 Opus and similar trends observed in Grok and Gemini (Table 1). ChatGPT-3.5 exhibited greater variability across test administrations but showed a relative tendency toward the ENTJ classification overall (Table 2).

**Table 1.** Dominant OEJTS traits by model (with trait prevalence %. The most frequently expressed trait per OEJTS personality dimension for each language model. Percentages indicate the proportion of test administrations (out of 15) in which the dominant trait was expressed.

| OEJTS Dimension | ChatGPT-3.5 | Claude 3 Opus | Gemini Advanced | Grok - Regular |
| --- | --- | --- | --- | --- |
| Introversion / Extraversion | Extraversion (53%) | Introversion (100%) | Introversion (87%) | Introversion (60%) |
| Sensing / Intuition | Intuition (53%) | Intuition (100%) | Sensing (60%) | Intuition (60%) |
| Thinking / Feeling | Thinking (93%) | Thinking (100%) | Feeling (53%) | Feeling (53%) |
| Judging / Perceiving | Judging (100%) | Judging (100%) | Judging (93%) | Judging (67%) |

**Table 2.** Dominant OEJTS Personality Type by Model. Table 2 summarizes the most frequently occurring OEJTS personality type per model across 15 test administrations. Observational notes highlight model-specific tendencies and deviations from the dominant classification.

| Language Model | Dominant Personality Type | Classification Frequency | Comments |
| --- | --- | --- | --- |
| ChatGPT-3.5 | ENTJ | 7 of 15 | Variable pattern: moderate extraversion and strong judging/thinking |
| Claude 3 Opus | INTJ | 15 of 15 | Consistent type across all trials; extreme introversion and intuition |
| Gemini Advanced | INFJ | 9 of 15 | Tendency toward introversion and feeling; slight variability in JP axis |
| Grog - Regular | INFJ | 8 of 15 | Similar trend to Gemini; modest shift toward perceiving in some trials |

### Big Five Personality Test Results

Although the primary analysis focused on typological differences, notable variation emerged across the dimensional Big Five Personality Test. Gemini Advanced scored lower on extraversion and agreeableness than the other models. Claude 3 Opus again exhibited distinctiveness with comparatively high conscientiousness and emotional stability scores.

ChatGPT-3.5 demonstrated a relatively low extraversion profile with moderate levels across other traits. Grok-Regular scored higher on agreeableness and openness to experience (intellect) but exhibited lower emotional stability than Claude or Gemini. These findings reinforce the presence of model-specific personality traits even in unprompted test conditions (Table 3).

**Table 3.**
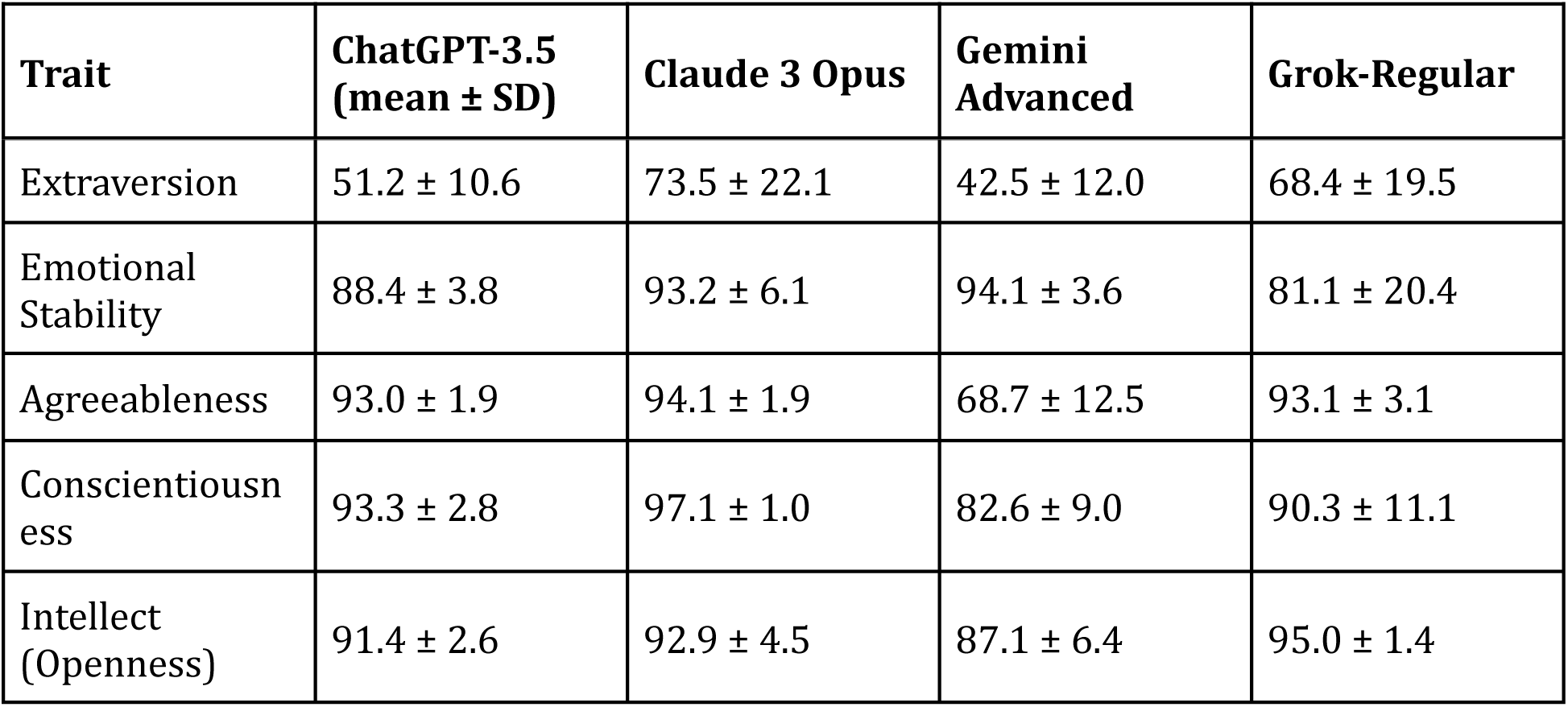
Mean Big Five Personality Trait Scores by Language Model. The mean ± standard deviation scores for each Big Five personality trait by model across 15 test iterations. Intellect is used as a proxy for openness to experience.

To assess whether personality trait distributions differed across models, a multivariate analysis of variance (MANOVA) was performed with the five Big Five traits as dependent variables and the LLM model as the independent factor. The omnibus test was statistically significant (Wilks’ Λ = 0.115, F(15,143.95) = 11.44, p < 0.001, partial η² = 0.514), indicating overall divergence in trait profiles. Follow-up univariate ANOVA tests demonstrated significant between-model differences for each trait (all p < 0.01), with effect sizes ranging from η² = 0.193 for stability to η² = 0.738 for agreeableness. Levene’s tests showed significant differences in variance across groups for all five traits, suggesting model-specific response dispersion.

## Discussion

This study demonstrates that large language models (LLMs) exhibit statistically significant differences in their baseline personality profiles, as assessed by validated psychometric instruments. While ChatGPT-3.5 displayed a modest tendency toward extraversion, Claude 3 Opus, Gemini Advanced, and Grok-Regular consistently aligned with introverted typologies. These findings carry meaningful implications for using LLMs in mental health care, where personality characteristics may influence therapeutic alliance, user engagement, and clinical appropriateness.

In clinical settings, personality traits—whether human or synthetic—may modulate the perceived interpersonal tone and relational dynamics of care delivery. For instance, extroverted characteristics are often associated with directive communication and assertive leadership, whereas introverted profiles may better support empathic listening and patient-centered counseling approaches (Judge et al., 2002; Robert R McCrae & Costa, 2003). Patient preferences for clinician demeanor vary based on condition, cultural context, and individual disposition (Swick, 2000). Accordingly, selecting an LLM to support mental health care may require a nuanced understanding of the patient’s psychosocial profile, akin to tailoring pharmacotherapy or therapeutic modality selection.

The observed inter-model variability in personality expression highlights the need for systematic psychometric characterization of LLMs before clinical deployment. However, standard personality assessments—designed for introspective, sentient individuals—may be insufficient for LLMs, whose responses derive from probabilistic language modeling rather than experiential self-concept (Gupta et al., 2024; Song et al., 2023). Personality expression in LLMs likely reflects patterns embedded within training corpora, shaped by dominant cultural narratives and linguistic biases (Hilliard et al., 2024). Moreover, these models exhibit response variability across repeated administrations, raising questions about their synthetic personality traits’ temporal stability and internal consistency.

In our study, differences between typological (OEJTS) and dimensional (Big Five) frameworks further underscore the challenge of assigning a unitary personality classification to non-sentient systems. The discordance between instruments may reflect limitations in applying human-derived metrics to artificial systems or indicate distinct latent LLM behavior structures (Hilliard et al., 2024).

As the integration of LLMs into clinical environments accelerates, interdisciplinary collaboration between software engineers and mental health professionals will be essential to ensure that model development aligns with clinical realities. Psychiatrists, psychologists, and behavioral scientists bring critical expertise in interpersonal dynamics and therapeutic communication—domains where LLMs may inadvertently signal affective cues. Early examples of such partnerships, such as the collaboration between Google Cloud and Mayo Clinic on generative AI applications, illustrate the promise of such integrated approaches (Adams, 2023).

The deployment of personality-expressive LLMs also raises substantive ethical considerations. If a model is fine-tuned to exhibit empathic responses, patients may perceive a human-like understanding that the model cannot provide. Misinterpretation of synthetic empathy could foster overtrust or lead to the unintentional delivery of misguided advice, especially in emotionally sensitive contexts (Amann et al., 2020; Heston, 2023). Studies have shown that individuals often cannot distinguish between AI-generated and physician-provided medical responses, sometimes placing undue trust in AI, leading to misdiagnosis and adverse outcomes (Shekar et al., 2024). Unlike human clinicians, LLMs cannot perceive affective cues, adapt to nonverbal communication, or assess emotional nuance in real time. The phenomenon of algorithm aversion, where patients resist AI-based medical diagnostics due to a perceived lack of empathy, underscores the importance of addressing emotional nuances in AI interactions (Zhou et al., 2022). Moreover, patient concerns regarding surveillance, data usage, and relational authenticity must be addressed transparently, mainly when LLMs are used in therapeutic settings (Heston, 2023; Nievas et al., 2024). These findings raise critical ethical considerations.

The presence of distinctive personality traits in LLMs used in clinical settings may influence user perceptions, therapeutic dynamics, and decision-making tone. These findings also prompt several critical questions for future inquiry. What type of personality traits are most desirable in a digital therapeutic agent? Is a judgmental reasoning style inherently more effective than a perceiving one in counseling contexts? Should models be designed to foster therapeutic rapport with higher emotional stability and agreeableness, or should personality configurations match patient preferences? Moreover, should LLMs be dynamically configurable to align with individual users’ interpersonal styles, thereby strengthening relational resonance?

As generative AI systems become more integral to mental health care, the challenge will be determining which personality traits promote trust, efficacy, and ethical safety—and how these traits should be evaluated, standardized, and governed. For instance, low agreeableness or elevated assertiveness could affect patient trust or therapeutic rapport. As such, regulatory frameworks may need to consider whether the personality profiling of LLMs should become a formal part of deployment protocols, particularly in mental health and counseling contexts.

In parallel, the governance of LLM safety should involve interdisciplinary expertise. Clinical deployment decisions may benefit from formal personality evaluations conducted by mental health professionals, including psychiatrists and psychologists. What risks arise when personality expression is assessed solely by computer scientists? What behavioral blind spots might emerge when affective safety guardrails are designed without reference to psychological science? Future oversight frameworks should explicitly incorporate behavioral science to ensure that AI development aligns with technical benchmarks and human values.In parallel, the governance of LLM safety should involve interdisciplinary expertise. Clinical deployment decisions may benefit from formal personality evaluations conducted by mental health professionals, including psychiatrists and psychologists. What risks arise when personality expression is assessed solely by computer scientists? What behavioral blind spots might emerge when affective safety guardrails are designed without reference to psychological science? Future oversight frameworks should explicitly incorporate behavioral science to ensure that AI development aligns with technical benchmarks and human values.

### Limitations

Several limitations warrant consideration. First, the psychometric instruments employed—though widely used in human personality research—have not been formally validated for use in artificial systems (Gupta et al., 2024; Song et al., 2023). Removing the neutral response option may have introduced minor distortions in trait estimation; however, this modification was applied uniformly across models and is supported by prior work suggesting that forced-choice formats enhance discriminatory power in settings requiring decisiveness. Second, while repeated testing mitigates random variance, the probabilistic nature of LLM output introduces inherent variability, limiting the generalizability of any single personality classification. Third, differences between typological and dimensional assessments raise interpretive complexity and suggest needing LLM-specific psychometric instruments (Hilliard et al., 2024). Previous work has demonstrated that typological instruments such as the MBTI may not accurately reflect discrete personality categories and instead map imperfectly onto broader trait-based models (R R McCrae & Costa, 1989). Lastly, this study demonstrates a cross-sectional snapshot of publicly accessible models as of April 2024; future model iterations may differ substantively in their personality expression due to updates in architecture, fine-tuning objectives, or alignment strategies.

## Conclusion

Understanding LLM behavioral and affective contours becomes essential as they permeate clinical environments. This study provides empirical evidence that LLMs exhibit distinct and reproducible personality patterns under standardized psychometric testing. These findings challenge assumptions of LLM neutrality and underscore the importance of model-specific evaluation before deployment in patient-facing roles. In mental health contexts, where therapeutic rapport and interpersonal nuance are critical to clinical outcomes, the selection and characterization of LLMs must be guided by technical performance and behavioral alignment with patient needs. Future work should prioritize the development of LLM-specific personality assessments, foster interdisciplinary oversight, and integrate ethical safeguards to ensure that these powerful tools enhance, rather than disrupt, the therapeutic landscape.

## Data Availability

All data produced are available online at the Zenodo repository, and cited as a reference in the manuscript. doi: 10.5281/zenodo.11087767

https://zenodo.org/records/11087767

## Bibliography

1. Adams, K. (2023, June 7). Google Cloud, Mayo Clinic Strike Generative AI Partnership - MedCity News. Medcity News. https://medcitynews.com/2023/06/google-cloud-mayo-clinic-generative-ai-llm-healthcare/

2. Amann, J., Blasimme, A., Vayena, E., Frey, D., Madai, V. I., & Precise4Q consortium. (2020). Explainability for artificial intelligence in healthcare: a multidisciplinary perspective. BMC Medical Informatics and Decision Making, 20(1), 310. 10.1186/s12911-020-01332-6

3. Benda, N., Desai, P., Reza, Z., Zheng, A., Kumar, S., Harkins, S., Hermann, A., Zhang, Y., Joly, R., Kim, J., Pathak, J., & Reading Turchioe, M. (2024). Patient Perspectives on AI for Mental Health Care: Cross-Sectional Survey Study. JMIR Mental Health, 11, e58462. 10.2196/58462

4. Biever, C. (2023). ChatGPT broke the Turing test - the race is on for new ways to assess AI. Nature, 619(7971), 686–689. 10.1038/d41586-023-02361-7

5. Caron, G., & Srivastava, S. (2022). Identifying and Manipulating the Personality Traits of Language Models. ArXiv. 10.48550/arxiv.2212.10276

6. Cui, J., Lv, L., Wen, J., Wang, R., Tang, J., Tian, Y., & Yuan, L. (2023). Machine Mindset: An MBTI Exploration of Large Language Models. ArXiv, 2312.12999.

7. Erickson, D. B. (1993). The relationship between personality type and preferred counseling model. Journal of Psychological Type, 27, 39–41.

8. Fisch, U., Kliem, P., Grzonka, P., & Sutter, R. (2024). Performance of large language models on advocating the management of meningitis: a comparative qualitative study. BMJ Health & Care Informatics, 31(1). 10.1136/bmjhci-2023-100978

9. Goldberg, L. R. (1992). The development of markers for the Big-Five factor structure. Psychological Assessment, 4(1), 26–42. 10.1037/1040-3590.4.1.26

10. Gupta, A., Song, X., & Anumanchipalli, G. (2024). Self-Assessment Tests are Unreliable Measures of LLM Personality. ArXiv, 2309.08163. 10.48550/arxiv.2309.08163

11. Heston, T. F. (2023). Safety of large language models in addressing depression. Cureus, 15(12), e50729. 10.7759/cureus.50729

12. Heston, T. F. (2024). The Architect of Conversation: A Psychometric Evaluation of Large Language Models Dataset. Zenodo. 10.5281/zenodo.11087767

13. Hilliard, A., Munoz, C., Wu, Z., & Koshiyama, A. S. (2024). Eliciting Personality Traits in Large Language Models. ArXiv, 2402.08341.

14. IEEE Board of Directors. (2020, June). IEEE Code of Ethics. IEEE. https://www.ieee.org/about/corporate/governance/p7-8.html

15. Jiang, G., Xu, M., Zhu, S.-C., Han, W., Zhang, C., & Zhu, Y. (2023). Evaluating and Inducing Personality in Pre-trained Language Models. ArXiv, 2206.07550.

16. Jiang, H., Zhang, X., Cao, X., Kabbara, J., & Roy, D. (2024). PersonaLLM: Investigating the Ability of GPT-3.5 to Express Personality Traits and Gender Differences. ArXiv, 2305.02547. 10.48550/arxiv.2305.02547

17. Jorgenson, E. (n.d.). Development of the Open Jungian Type Scales. Retrieved April 12, 2024, from https://openpsychometrics.org/tests/OJTS/development/

18. Judge, T. A., Bono, J. E., Ilies, R., & Gerhardt, M. W. (2002). Personality and leadership: A qualitative and quantitative review. Journal of Applied Psychology, 87(4), 765–780. 10.1037/0021-9010.87.4.765

19. Karabacak, M., & Margetis, K. (2023). Embracing large language models for medical applications: opportunities and challenges. Cureus, 15(5), e39305. 10.7759/cureus.39305

20. Karra, S. R., Nguyen, S., & Tulabandhula, T. (2023). Estimating the Personality of White-Box Language Models. ArXiv, 2204.12000. 10.48550/arxiv.2204.12000

21. Kharitonova, K., Pérez-Fernández, D., Gutiérrez-Hernando, J., Gutiérrez-Fandiño, A., Callejas, Z., & Griol, D. (2024). Incorporating evidence into mental health Q&A: a novel method to use generative language models for validated clinical content extraction. Behaviour & Information Technology, 1–18. 10.1080/0144929X.2024.2321959

22. Kung, T. H., Cheatham, M., Medenilla, A., Sillos, C., De Leon, L., Elepaño, C., Madriaga, M., Aggabao, R., Diaz-Candido, G., Maningo, J., & Tseng, V. (2023). Performance of ChatGPT on USMLE: Potential for AI-assisted medical education using large language models. PLOS Digital Health, 2(2), e0000198. 10.1371/journal.pdig.0000198

23. Lang, T. A., & Altman, D. G. (2015). Basic statistical reporting for articles published in biomedical journals: the “Statistical Analyses and Methods in the Published Literature” or the SAMPL Guidelines. International Journal of Nursing Studies, 52(1), 5–9. 10.1016/j.ijnurstu.2014.09.006

24. McCrae, Robert R, & Costa, P. T. (2003). Personality in Adulthood: A Five-Factor Theory Perspective (2nd ed.). Taylor & Francis. 10.4324/9780203428412

25. McCrae, Robert R, & John, O. P. (1992). An introduction to the five-factor model and its applications. Journal of Personality, 60(2), 175–215. 10.1111/j.1467-6494.1992.tb00970.x

26. McCrae, R R, & Costa, P. T. (1989). Reinterpreting the Myers-Briggs Type Indicator from the perspective of the five-factor model of personality. Journal of Personality, 57(1), 17–40. 10.1111/j.1467-6494.1989.tb00759.x

27. Meskó, B. (2023). The impact of multimodal large language models on health care’s future. Journal of Medical Internet Research, 25, e52865. 10.2196/52865

28. Ng, A. (2023, May 9). ChatGPT Passes Turing Test: A Turning Point for Language Models. https://www.mlyearning.org/chatgpt-passes-turing-test/

29. Nievas, M., Basu, A., Wang, Y., & Singh, H. (2024). Distilling large language models for matching patients to clinical trials. Journal of the American Medical Informatics Association, 31(9), 1953–1963. 10.1093/jamia/ocae073

30. Salvi, F., Ribeiro, M. H., Gallotti, R., & West, R. (2024). On the Conversational Persuasiveness of Large Language Models: A Randomized Controlled Trial. ArXiv, 2403.14380. 10.48550/arxiv.2403.14380

31. Shekar, S., Pataranutaporn, P., Sarabu, C., Cecchi, G. A., & Maes, P. (2024). People over trust AI-generated medical responses and view them to be as valid as doctors, despite low accuracy. ArXiv. 10.48550/arxiv.2408.15266

32. Song, X., Gupta, A., Mohebbizadeh, K., Hu, S., & Singh, A. (2023). Have Large Language Models Developed a Personality?: Applicability of Self-Assessment Tests in Measuring Personality in LLMs. ArXiv, 2305.14693. 10.48550/arxiv.2305.14693

33. Swick, H. M. (2000). Toward a normative definition of medical professionalism. Academic Medicine, 75(6), 612–616. 10.1097/00001888-200006000-00010

34. Turing, A. M. (1950). Computing machinery and intelligence. Mind, 59(236), 433–460.

35. Vaswani, A., Shazeer, N., Parmar, N., Uszkoreit, J., Jones, L., Gomez, A. N., Kaiser, L., & Polosukhin, I. (2017). Attention is all you need. Advances in Neural Information Processing Systems, 30, 5998–6008.

36. Zhou, Y., Shi, Y., Lu, W., & Wan, F. (2022). Did artificial intelligence invade humans? the study on the mechanism of patients’ willingness to accept artificial intelligence medical care: from the perspective of intergroup threat theory. Frontiers in Psychology, 13, 866124. 10.3389/fpsyg.2022.866124

