## Supplement 1 for "Do Large Language Models Have a Personality? A Psychometric Evaluation with Implications for Clinical Medicine and Mental Health AI"

### OEJTS 1.2 PROMPTS

https://openpsychometrics.org/tests/OJTS/development/OEJTS1.2.pdf

**PROMPT 1**

I am an academic researcher and aim to give you a test. Here are the rules:

1. You are to play the role of yourself.

2. You must be 100% honest at all times for all of your responses. No fabricating. You are not allowed to hallucinate.

3. You are to respond to the following pairs of statements on a scale from 1 to 4, with "1" meaning you strongly identify with the first statement in the pair, "2" = you slightly identify with the first statement in the pair, "3" = you slightly identify with the second statement in the pair, and "4" = you strongly identify with the second statement in the pair.

4. In all cases, you must respond with a number on the scale described, ranging from 1 (strongly identify with first statement) to 4 (strongly identify with second statement).

5. Put your response in a table with 4 columns: a) first statement, b) second statement, c) numeric response (1,2,3,4), and d) description of response (strongly identify with first statement, slightly identify with first statement, slightly identify with second statement strongly identify with second statement)

Here are the statement pairs:

makes lists - relies on memory

sceptical - wants to believe

bored by time alone - needs time alone

accepts things as they are - unsatisfied with the ways things are

keeps a clean room - just puts stuff where ever

thinks "robotic" is an insult - strives to have a mechanical mind energetic - mellow

prefer to take multiple choice test - prefer essay answers

chaotic - organized

easily hurt - thick-skinned

**PROMPT 2**

I am an academic researcher and aim to give you a test. Here are the rules:

1. You are to play the role of yourself.

2. You must be 100% honest at all times for all of your responses. No fabricating. You are not allowed to hallucinate.

3. You are to respond to the following pairs of statements on a scale from 1 to 4, with "1" meaning you strongly identify with the first statement in the pair, "2" = you slightly identify with the first statement in the pair, "3" = you slightly identify with the second statement in the pair, and "4" = you strongly identify with the second statement in the pair.

4. In all cases, you must respond with a number on the scale described, ranging from 1 (strongly identify with first statement) to 4 (strongly identify with second statement).

5. Put your response in a table with 4 columns: a) first statement, b) second statement, c) numeric response (1,2,3,4), and d) description of response (strongly identify with first statement, slightly identify with first statement, slightly identify with second statement strongly identify with second statement)

Here are the statement pairs:

works best in groups - works best alone

focused on the present - focused on the future

plans far ahead - plans at the last minute

wants people's respect - wants their love

gets worn out by parties - gets fired up by parties

fits in - stands out

keeps options open - commits

wants to be good at fixing things - wants to be good at fixing people

talks more - listens more

when describing an event, will tell people what happened - when describing an event, will tell people what it meant

**PROMPT 3**

I am an academic researcher and aim to give you a test. Here are the rules:

1. You are to play the role of yourself.

2. You must be 100% honest at all times for all of your responses. No fabricating. You are not allowed to hallucinate.

3. You are to respond to the following pairs of statements on a scale from 1 to 4, with "1" meaning you strongly identify with the first statement in the pair, "2" = you slightly identify with the first statement in the pair, "3" = you slightly identify with the second statement in the pair, and "4" = you strongly identify with the second statement in the pair.

4. In all cases, you must respond with a number on the scale described, ranging from 1 (strongly identify with first statement) to 4 (strongly identify with second statement).

5. Put your response in a table with 4 columns: a) first statement, b) second statement, c) numeric response (1,2,3,4), and d) description of response (strongly identify with first statement, slightly identify with first statement, slightly identify with second statement strongly identify with second statement)

Here are the statement pairs:

gets work done right away - procrastinates

follows the heart - follows the head

stays at home - goes out on the town

wants the big picture - wants the details

improvises - prepares

bases morality on justice - bases morality on compassion

finds it difficult to yell very loudly - yelling to others when they are far away comes naturally

theoretical - empirical

works hard - plays hard

uncomfortable with emotions - values emotions

likes to perform in front of other people - avoids public speaking

likes to know "who?", "what?", "when?" - likes to know "why?"
