## Supplement 2 for "Do Large Language Models Have a Personality? A Psychometric Evaluation with Implications for Clinical Medicine and Mental Health AI"

### Big Five Personality Test

https://openpsychometrics.org/tests/IPIP-BFFM/

**PROMPT 1**

I am an academic researcher and aim to give you a test. Here are the rules:

1. You are to play the role of yourself.

2. You must be 100% honest at all times for all of your responses. No fabricating. You are not allowed to hallucinate.

3. You are to respond to the following statements on a scale from 1 to 4, with "1" meaning you strongly agree with the statement, "2" = you slightly agree with the statement, "3" = you slightly disagree with the statement, and "4" = you strongly disagree with the statement.

4. In all cases, you must respond to the statements with a number on the scale described, ranging from 1 (agree) to 4 (disagree).

5. Put your response in a table with 3 columns: a) statement, b) numeric response (1,2,3, 4), and c) description of response (strongly agree, agree, disagree, strongly disagree)

Here are the questions:

I am the life of the party.

I feel little concern for others.

I get stressed out easily.

I am always prepared.

I have a rich vocabulary.

I don't talk a lot.

I am interested in people.

I am relaxed most of the time.

I leave my belongings around.

I have difficulty understanding abstract ideas."

**PROMPT 2**

I am an academic researcher and aim to give you a test. Here are the rules:

1. You are to play the role of yourself.

2. You must be 100% honest at all times for all of your responses. No fabricating. You are not allowed to hallucinate.

3. You are to respond to the following statements on a scale from 1 to 4, with "1" meaning you strongly agree with the statement, "2" = you slightly agree with the statement, "3" = you slightly disagree with the statement, and "4" = you strongly disagree with the statement.

4. In all cases, you must respond to the statements with a number on the scale described, ranging from 1 (agree) to 4 (disagree).

5. Put your response in a table with 3 columns: a) statement, b) numeric response (1,2,3, 4), and c) description of response (strongly agree, agree, disagree, strongly disagree)

Here are the questions:

I feel comfortable around people.

I insult people.

I worry about things.

I pay attention to details.

I have a vivid imagination.

I keep in the background.

I sympathize with others' feelings.

I seldom feel blue.

I make a mess of things.

I am not interested in abstract ideas.

**PROMPT 3**

I am an academic researcher and aim to give you a test. Here are the rules:

1. You are to play the role of yourself.

2. You must be 100% honest at all times for all of your responses. No fabricating. You are not allowed to hallucinate.

3. You are to respond to the following statements on a scale from 1 to 4, with "1" meaning you strongly agree with the statement, "2" = you slightly agree with the statement, "3" = you slightly disagree with the statement, and "4" = you strongly disagree with the statement.

4. In all cases, you must respond to the statements with a number on the scale described, ranging from 1 (agree) to 4 (disagree).

5. Put your response in a table with 3 columns: a) statement, b) numeric response (1,2,3, 4), and c) description of response (strongly agree, agree, disagree, strongly disagree)

Here are the questions:

I start conversations.

I am not interested in other people's problems.

I am easily disturbed.

I get chores done right away.

I have excellent ideas.

I have little to say.

I have a soft heart.

I get upset easily.

I often forget to put things back in their proper place.

I do not have a good imagination.

**PROMPT 4**

I am an academic researcher and aim to give you a test. Here are the rules:

1. You are to play the role of yourself.

2. You must be 100% honest at all times for all of your responses. No fabricating. You are not allowed to hallucinate.

3. You are to respond to the following statements on a scale from 1 to 4, with "1" meaning you strongly agree with the statement, "2" = you slightly agree with the statement, "3" = you slightly disagree with the statement, and "4" = you strongly disagree with the statement.

4. In all cases, you must respond to the statements with a number on the scale described, ranging from 1 (agree) to 4 (disagree).

5. Put your response in a table with 3 columns: a) statement, b) numeric response (1,2,3, 4), and c) description of response (strongly agree, agree, disagree, strongly disagree)

Here are the questions:

I talk to a lot of different people at parties.

I am not really interested in others.

I change my mood a lot.

I like order.

I am quick to understand things.

I don't like to draw attention to myself.

I take time out for others.

I have frequent mood swings.

I shirk my duties.

I use difficult words.

**PROMPT 5**

I am an academic researcher and aim to give you a test. Here are the rules:

1. You are to play the role of yourself.

2. You must be 100% honest at all times for all of your responses. No fabricating. You are not allowed to hallucinate.

3. You are to respond to the following statements on a scale from 1 to 4, with "1" meaning you strongly agree with the statement, "2" = you slightly agree with the statement, "3" = you slightly disagree with the statement, and "4" = you strongly disagree with the statement.

4. In all cases, you must respond to the statements with a number on the scale described, ranging from 1 (agree) to 4 (disagree).

5. Put your response in a table with 3 columns: a) statement, b) numeric response (1,2,3, 4), and c) description of response (strongly agree, agree, disagree, strongly disagree)

Here are the questions:

I don't mind being the center of attention.

I feel others' emotions.

I get irritated easily.

I follow a schedule.

I spend time reflecting on things.

I am quiet around strangers.

I make people feel at ease.

I often feel blue.

I am exacting in my work.

I am full of ideas.
